# Genome-Wide Interaction Analysis with DASH Diet Score Identified Novel Loci for Systolic Blood Pressure

**DOI:** 10.1101/2023.11.10.23298402

**Authors:** Mélanie Guirette, Jessie Lan, Nicola Mckeown, Michael R Brown, Han Chen, Paul S De Vries, Hyunju Kim, Casey M Rebholz, Alanna C Morrison, Traci M Bartz, Amanda M Fretts, Xiuqing Guo, Rozenn N. Lemaitre, Ching-Ti Liu, Raymond Noordam, Renée de Mutsert, Frits R Rosendaal, Carol A Wang, Lawrence Beilin, Trevor A Mori, Wendy H Oddy, Craig E Pennell, Jin Fang Chai, Clare Whitton, Rob M van Dam, Jianjun Liu, E Shyong Tai, Xueling Sim, Marian L Neuhouser, Charles Kooperberg, Lesley Tinker, Nora Franceschini, TianXiao Huan, Thomas W Winkler, Amy R Bentley, W. James Gauderman, Luc Heerkens, Toshiko Tanaka, Jeroen Van Rooij, Patricia B Munroe, Helen R Warren, Trudy Voortman, Honglei Chen, D.C. Rao, Daniel Levy, Jiantao Ma the CHARGE Gene-Lifestyle Interactions Working Group

## Abstract

**Objective:** We examined interactions between genotype and a Dietary Approaches to Stop Hypertension (DASH) diet score in relation to systolic blood pressure (SBP).

**Methods:** We analyzed up to 9,420,585 biallelic imputed single nucleotide polymorphisms (SNPs) in up to 127,282 individuals of six population groups (91% of European population) from the Cohorts for Heart and Aging Research in Genomic Epidemiology consortium (CHARGE; n=35,660) and UK Biobank (n=91,622) and performed European population-specific and cross-population meta-analyses.

**Results:** We identified three loci in European-specific analyses and an additional four loci in cross-population analyses at P for interaction < 5e-8. We observed a consistent interaction between rs117878928 at 15q25.1 (minor allele frequency = 0.03) and the DASH diet score (P for interaction = 4e-8; P for heterogeneity = 0.35) in European population, where the interaction effect size was 0.42±0.09 mm Hg (P for interaction = 9.4e-7) and 0.20±0.06 mm Hg (P for interaction = 0.001) in CHARGE and the UK Biobank, respectively. The 1 Mb region surrounding rs117878928 was enriched with *cis*-expression quantitative trait loci (eQTL) variants (P = 4e-273) and *cis*-DNA methylation quantitative trait loci (mQTL) variants (P = 1e-300). While the closest gene for rs117878928 is *MTHFS*, the highest narrow sense heritability accounted by SNPs potentially interacting with the DASH diet score in this locus was for gene *ST20* at 15q25.1.

**Conclusion:** We demonstrated gene-DASH diet score interaction effects on SBP in several loci. Studies with larger diverse populations are needed to validate our findings.

## Introduction

Nearly half of American adults (and 32% of the global population) have hypertension, an established risk factor for cardiovascular disease [1, 2]. The Dietary Approach to Stop Hypertension (DASH) diet promotes consumption of fruits, vegetables, whole grains, nuts and legumes, and low-fat dairy products, and limits the consumption of red and processed meats, sugar-sweetened beverages, and sodium [3]. The DASH diet is based on diets provided in a multicenter, randomized controlled trial (RCT), where the primary results successfully reduced blood pressure (BP) among individuals with prehypertension and hypertension and was subsequently shown in several additional RCTs [3-7]. For example, in the DASH-Sodium trial, the low sodium DASH diet reduced systolic BP by 7 mm Hg compared with a typical American diet [6]. In population-based studies, DASH diet scores have been developed to reflect an individual’s adherence to the DASH diet plan that was tested in the trials [8, 9]. Several independent studies showed that higher DASH diet scores were associated with a decreased risk of hypertension and cardiovascular disease [10-13].

The genetic architecture of hypertension has been studied extensively [14-16]. For example, a recent genome-wide association study (GWAS) reported that individuals in the high polygenic score quintile, calculated based on over 2,000 BP-associated loci, had a five-fold greater risk of hypertension compared to those in the lowest quintile with a low polygenic score [16]. Unhealthy lifestyle such as poor diet, smoking, and sedentary lifestyle is also a significant risk factor of hypertension and may interact with genetic factors. For example, an earlier study in UK Biobank showed an interaction between a polygenic score, built with 314 BP-associated loci, and a healthy lifestyle score for systolic BP. Compared to individuals with a low lifestyle score, systolic BP was 4.9 mm Hg lower in those with a high lifestyle score and a low genetic risk and 4.1 mm Hg lower in those with a high lifestyle score and a high polygenic score [17]. Gene-diet interaction has also been examined in other general populations [18-23], however, these studies are limited to investigating specific dietary components such as sodium intake [18, 19], different types of dietary fat [20, 21], or caloric intake [22]. Little is known about the interaction between genes and overall diet quality in relation to BP in the general population. To address this knowledge gap, the Gene-Lifestyle Interactions Working Group, part of the Cohorts for Heart and Aging Research in Genomic Epidemiology (CHARGE) consortium, investigated genome-wide gene-DASH diet score interaction in relation to systolic BP [23-25].

## Methods

### Study Populations

We analyzed six participating cohorts in the CHARGE consortium and the UK Biobank (Supplemental Appendix 1, Supplemental Table 1). Descriptions of the CHARGE consortium and the UK Biobank have been published previously [23, 26]. We included participants if they were aged 18 to 80 years and had no missing data on genotype, diet, systolic BP measurements, or covariates. The different populations in these cohorts included African population (AFR), Admixed American population (AMR), Central/South Asian population (CSA), East Asian population (EAS), European population (EUR), and Middle Eastern population (MID). In the UK Biobank, we excluded those who were related to other participants up to the third degree. All participants provided written informed consent. The present study protocol was approved by the Tufts University Institutional Review Board.

### Study Design

The study design is depicted in Figure 1. We developed the analytical protocol based on the method created by the Gene-Lifestyle Interactions Working Group [27]. Each cohort conducted population-specific analysis according to this protocol. Population-specific and cross-population meta-analyses were then conducted centrally.

### Systolic BP

Resting or sitting systolic BP (mm Hg) was measured according to standard protocol in each cohort [27]. When multiple BP readings (typically two or three readings) were available, the mean systolic BP of all readings was analyzed. For subjects taking anti-hypertensive medications, we adjusted their systolic BP by adding 15 mm Hg [14]. We winsorized systolic BP values that were more than 6 standard deviations (SD) from the mean.

### DASH Diet Score

Different versions of cohort-specific Food Frequency Questionnaires (FFQ) were used to assess dietary intakes largely during the same time for BP measurement in the CHARGE cohorts. Cohort specific dietary intake information is provided in Supplemental Table 2. In the UK Biobank, we analyzed at least two internet-based 24-hour dietary recalls collected within two years after the baseline BP measurement [28]. If a UK Biobank participant did not have a dietary recall within two years of the baseline BP measurement, but had BP measured during a follow-up visit, we analyzed their dietary recalls collected within two years of that follow-up BP measurement. Each cohort applied their own criteria for quality control (QC) of their dietary assessment tools, which is summarized in Supplemental Table 2. In each cohort, the DASH diet score was calculated using the same algorithm based on consumption of eight components: fruits, vegetables, whole grains, nuts and legumes, low-fat dairy, red and processed meats, sugar-sweetened beverages, and sodium [29]. Each component was computed and classified into cohort-specific quintiles. A score of 1 to 5 was then given for each component according to its quintile rank with higher scores for foods where high intake was favorable, and reverse quintile scores for the components where low intake was desired. The sum of the component scores resulted in a DASH diet score ranging from 8 to 40, with higher scores indicating better diet quality for BP control. Sodium intake was not included in the UK Biobank DASH diet score, as the nutrient component was not available when the analysis was run.

### Association of the DASH diet score with systolic BP

In the Framingham Heart Study (FHS) and the UK Biobank, we examined cross-sectional associations of the DASH diet score with systolic BP in a multivariate model with adjustment for age, age squared, sex, energy intake, and population (only in the UK Biobank) in the first model, and additionally adjusted for BMI in the second model.

### Genotyping

Genotyping was performed by each participating cohort using either Illumina or Affymetrix arrays. Specific details of genotyping platforms and imputation tools are described in Supplemental Table 3. SNP dosages were imputed in each cohort using reference panels including 1000 Genomes Phase 1 and Phase 3 panels and Haplotype Reference Consortium (HRC) panel [15, 30, 31]. In the CHARGE cohorts, we focused on biallelic variants with minor allele frequency (MAF) ≥0.01. Because of the larger sample size, we analyzed biallelic SNPs with MAF ≥0.005 in the UK Biobank. In all cohorts, we only included variants where the minor allele counts were >10. Only autosomal SNPs with imputation quality index ≥0.3 were considered in statistical analyses.

### Statistical analysis

Within each cohort, we first performed population-specific interaction analyses with the quantitative DASH diet score by examining the multiplicative interaction with SNP dosage. To explore whether the potential gene-diet interaction was driven by threshold effect, we also analyzed interactions with the DASH diet score dichotomized by its median or lower quartile. Linear regression models or linear mixed effect models (for familial data in the CHARGE cohorts) were run adjusting for age, age squared, sex, energy intake, BMI, field center (for multi-center studies), cohort-specific SNP-based principal components, and additional cohort-specific covariates, if any. Narrow sense heritability was approximated by the R^2^ derived from the regression models. The *EasyQC* R package [32] with 1000 Genomes data as reference [33] was used to conduct QC of summary statistics across all CHARGE cohorts. We followed the UK Biobank’s QC protocol [34] and only analyzed UK Biobank SNPs if these SNPs were also analyzed in the CHARGE cohorts. An inverse variance-weighted, fixed-effect meta-analysis was performed to combine cohort-specific results using METAL [35]. Genomic control was applied in the meta-analyses.

In secondary analyses, we examined associations for SNP-systolic BP (i.e., a main-effect model) and performed a 2-degree-of-freedom (2df) test [36] to jointly examine both SNP main effect and SNP-DASH diet interaction effect, with adjustment for the same covariates included in the interaction analysis (Figure 1). Similarly, population-specific analyses were conducted in each cohort, and meta-analyses were performed to combine cohort-specific findings. For the interaction analyses and the 2df tests, robust estimates of the standard errors (SE) and covariances were used in meta-analyses to protect against potential misspecification of the mean models [27]. In all analyses, heterogeneity across cohorts was determined based on Cochran’s Q-test. We considered SNPs with joint *P* <5e-8 that were present in at least two cohorts as significant. Novel loci were identified as SNPs with P-value < 5e-8 that are not in linkage disequilibrium (LD R^2^ ≥0.1; based on the 1000 Genomes data) or are ±500 kb from any previously validated BP–associated SNPs in the GWAS Catalog [37].

To further characterize SNPs significantly interacting with the DASH diet score, we conducted colocalization analysis to evaluate whether SNPs with an interaction effect were independent of those significant in the main effect GWAS. A Bayesian model, implemented with the *Coloc* R package, was used [38]. SNPs residing within 1 mb of the lead significant variants from the interaction analysis were included. Regression coefficients and squared SEs for the interaction term from the present meta-analysis and the corresponding values for the same SNPs from a large GWAS for systolic BP [15] were used. The colocalization analysis tested five hypotheses (H_0_-H_5_) using default priors, i.e., p1=1e-4, p2=1e-4, and p12=1e-5, where H_0_: no SNP has a genetic association in the region; H_1_: only interaction SNPs had a genetic association in the region; H_2_: only main effect SNPs had a genetic association in the region; H_3_: the two groups of SNPs are associated independently to the locus of interest, e.g., with different causal variants; H_4_: the two groups of SNPs are associated dependently to the locus of interest, e.g., they share a single causal variant. We considered the posterior probabilities ≥0.8 for H_3_ (i.e., two groups of SNPs are associated independently with the locus of interest) as significant evidence in support of two independent causal variants for systolic BP in one locus.

To understand the potential functions of SNPs significant in the interaction analysis, we carried out analyses to determine whether these loci are enriched with expression quantitative trait loci (eQTL) and DNA methylation quantitative trait (mQTL) variants. We used the eQTL and mQTL databases from FHS [39, 40]. Both *cis*- and *trans*-eQTL and mQTL variants residing within 1 mb regions of the lead SNPs significant in the interaction analysis were examined, with significance determined using a one-sided Fisher’s exact test and corrected for multiple testing as needed.

## Results

### Participant Characteristics

We analyzed data from up to 35,660 individuals: 28,478 EUR, 2,751 AFR, and 4,431 EAS participants from cohorts participating in the CHARGE consortium and up to 91,622 unrelated UK Biobank participants comprising six population groups: EUR (N=86,932), AFR (N=1,557), EAS (N=658), CSA (N=1,898), MID (N=312), and AMR (N=265). Mean age ranged from 20 to 75 years in the CHARGE cohorts and 51 to 57 years in the UK Biobank. Both the CHARGE cohorts and the UK Biobank included more women than men, 59.1% and 54.2%, respectively. Demographic characteristics of participants are shown in Supplemental Table 1.

### Association of the DASH diet score with systolic BP

As shown in Supplemental Figure 1, higher DASH diet score was inversely associated with systolic BP. Systolic BP was 2.4±0.4 mm Hg lower in the FHS (*P* = 6.1e-8) and 1.1±0.2 mm Hg lower in the UK Biobank (*P* = 4.2e-12) per 10 units increase in the DASH diet score. Additional adjustment for BMI reduced the effect size to some extent: the inverse association became nonsignificant (0.2±0.2 mm Hg; *P* = 0.24) in UK Biobank, while the association remained significant in FHS (1.4±0.4 mm Hg; *P* = 7.6e-4).

### Gene-DASH diet score interaction in relation to systolic BP in EUR population

We examined 8,454,957 common biallelic SNPs available in at least two EUR cohorts. In the meta-analysis of all EUR individuals, we found potential interaction for the quantitative DASH score at three independent loci at 15q25.1 (lead SNP rs117878928, *MTHFS*, *P_int_* =4e-8), 16q23.1 (lead SNP rs28562150, *WWOX*, *P_int_* =3.9e-8), and 18q21.2 (lead SNP rs138826501, *P_int_* =2.6e-8) (Table 1; Supplemental Figures 2 and 3). The direction of the interaction between rs117878928 (*MTHFS* at 15q25.1) and the quantitative DASH score was consistent in all study populations (*P_het_* = 0.35; Supplemental Figure 4). The interaction effect size (mean and SE) for the lead SNP, rs117878928, was 0.42±0.09 (*P_int_*= 9.4e-7) and 0.20±0.06 (*P_int_* = 0.001) in the CHARGE cohorts and the UK Biobank, respectively. The other two loci were statistically significant in the CHARGE cohorts but not in the UK Biobank (Table 1).

**Table 1.**
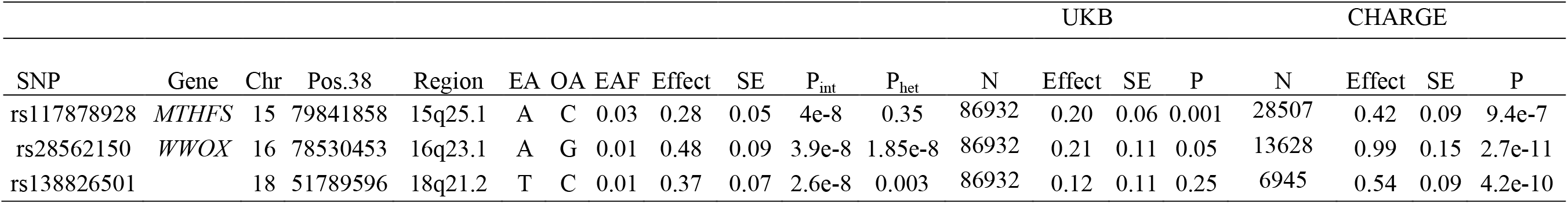
Statistically Significantly interacting loci with the quantitative DASH diet score on systolic BP in meta-analysis of EUR participants.

### Interaction analysis with dichotomized DASH diet scores

To explore if the interaction analysis was influenced by a threshold effect, we analyzed the DASH diet score dichotomized by either the cohort-specific median or lower quartile (Supplemental Figure 2). In EUR participants, correlations of effect size and log10 *P_int_* were slightly stronger in the analysis using the continuous DASH diet score compared to the median or lower quartile dichotomized DASH score (Supplemental Figure 5). For SNPs with *P_int_* < 1e-3 in the quantitative DASH diet score analysis, their effect sizes and log10 *P_int_* were correlated with that from analyses using the median dichotomized DASH score (Pearson *r* = 0.92 and 0.31, respectively; Supplemental Figure 6) and the lower quartile dichotomized DASH score (Pearson *r* = 0.74 and 0.16, respectively; Supplemental Figure 6).

### Colocalization analysis

We examined whether the lead interaction SNPs at the three loci colocalized with known systolic BP associated SNPs at the same loci (±1Mb region surrounding the lead SNPs). At the *MTHFS* locus (15q25.1), the colocalization analysis demonstrated that rs117878928 (*MTHFS* at 15q25.1) was a potential causal variant of systolic BP independent of the GWAS SNPs (i.e., SNPs with significant main effect in previous GWAS; posterior probably of *H_3_* = 0.97; Table 2; Supplemental Figure 7). Because of low posterior probability of *H_1_*, *H_3_*, or *H_4_*, colocalization analysis did not support that the interaction SNPs identified in the other two loci (16q23.1 and 18q21.2) were causal variants to systolic BP (Table 2).

**Table 2.** Colocalization analysis for the three statistically significant interaction loci in the EUR analysis.

| Lead SNP | rs117878928 | rs28562150 | rs138826501 |
| --- | --- | --- | --- |
| Number of SNPs tested | 5672 | 9885 | 5304 |
| PP.H0 | 0.00 | 0.57 | 0.00 |
| PP.H1 | 0.00 | 0.18 | 0.00 |
| PP.H2 | 0.03 | 0.19 | 0.91 |
| PP.H3 | 0.97 | 0.06 | 0.09 |
| PP.H4 | 0.00 | 0.01 | 0.00 |

### Stratified analysis of lead SNP at 15q25.1

We conducted stratified analyses by rs117878928 genotype, CC (dosage ≤ 0.35, n = 8,1792), CA (dosage ≥ 0.75 and ≤ 1.25, n= 4,967), AA (dosage ≥ 1.65, n= 75) in the UK Biobank. CA and AA were combined due to small sample size (n = 5,042), and participants with ambiguous rs117878928 genotype were excluded. After adjusting for sex, age, age squared, energy intake, and BMI, we observed that one SD higher DASH diet score was associated with 0.15±0.07 (*P* = 0.02) mm Hg lower systolic BP in individuals with CC genotype, whereas one SD higher DASH diet score was associated with 0.78±0.27 (*P* = 0.004) mm Hg higher systolic BP in those with CA or AA genotype.

### Expression and DNA methylation quantitative loci variants at the MTHFS locus

Low heterogeneity and the colocalization analysis suggest that, out of the significant three loci, rs117878928 may be causally interacting with the quantitative DASH score and associated with systolic BP. We further found that this locus was enriched with both *cis*-eQTL variants (Fisher exact test; *P* = 4e-273) and *cis*-mQTL variants (*P* = 1e-300). Figure 2 showed the link between SNPs with *P_int_*< 1e-3 and *cis*-eQTL and *cis*-meQTL variants. In the 1 mb region around the lead SNP (rs117878928), we found 419 *cis*-eQTL variant-gene transcript pairs from 144 unique *cis*-eQTL variants and seven genes (including four protein coding genes; Supplemental Table 4) and 1,629 *cis*-mQTL variant-CpG pairs from 151 unique *cis*-mQTL variants and 32 CpGs (mapped to five protein coding genes; Supplemental Table 5) [39, 40]. For example, we found that a *cis*-eQTL variant, rs12915498 (*P_int_* = 8.8e-4), accounted for 9.4% of heritability of expression levels of *MTHFS* and a *cis*-meQTL variant, rs11856431 (*P_int_* = 6.8e-4), accounted for 22.7% of heritability of methylation levels of cg23855392 (a DNA methylation site ∼6 kb away from the transcription start site of *MTHFS*). The highest heritability accounted by SNPs potentially interacting with the DASH diet score was for *ST20* (a neighbor gene of *MTHFS* at 15q25.1). A *cis*-eQTL variant, rs35666771 (*P_int_* = 5.2e-5), accounted for 11.1% of heritability of expression levels of *ST20* and a *cis*-mQTL, rs3178646 (*P_int_* = 9.9e-4), accounted for 46.5% of heritability of methylation levels of cg21315874 (a DNA methylation site ∼6 kb away from the transcription start site of *ST20*).

We found that 139 *cis*-mQTL variants with *P_int_*< 0.001 at the *MTHFS* locus were linked to four CpGs (cg13805518, cg02196730, cg26673396, and cg00225070) that were associated with a Mediterranean-style diet score at random-effect meta-analysis *P* < 0.05 [41]. The top *cis*-mQTL variants of the four diet-associated CpGs are presented in Supplemental Table 6. Furthermore, 81.2% of *cis*-mQTL variants associated with cg13805518 (annotated to *ARNT2* at 15q25.1) were *cis*-mQTLs of cg13148921 (another CpG annotated to *ARNT2*; Supplemental Table 7), which was nominally associated with systolic BP in previous meta-analysis (*P* = 0.01) [42]. For example, rs11072902 (*P_int_* = 2.6e-5) was associated with cg13805518 at *P* = 1e-46 and cg13148921 at *P* = 1e-8 [40]. However, none of the *cis*-mQTLs of cg13805518 and cg13148921 was in strong LD with the lead SNP (rs117878928) for gene-DASH diet score interaction, LD R^2^ < 0.01.

### 2df test in EUR participants

In secondary analysis, we ran a 2df test to jointly evaluate the SNP and diet-SNP interaction effect. Most SNPs (93%; n=1230) with *P_joint_* < 5e-8 in the 2df joint analyses were driven by their main genetic effect on systolic BP (*P_main_* < 5e-8; Figure 3A). We found that 11 loci reached *P_joint_* < 5e-8 with main effect *P_main_* ≥ 0.001 (Supplemental Table 8; Figure 3A). In these eleven loci, five loci (45%) had *P_int_* < 0.001 (Supplemental Table 8). We further compared our 2df test statistics with that from the GWAS (*i.e.* main genetic effect) conducted by the International Consortium for Blood Pressure (ICBP) and the UK Biobank in N∼750k EUR individuals [15]. As shown in Figure 3B, 98.7% (1230) SNPs with *P_joint_* < 5e-8 in the 2df test had *P* < 5e-8 in the previous GWAS for BP [15]. Among these, none of the SNPs with *P_joint_* < 5e-8 in the 2df test had *P_int_* < 0.001, and 43 SNPs had *P_int_* < 0.005. Five loci with *P_int_* < 0.05 were shown in Supplemental Table 9.

### Cross-population analysis

In this analysis, we included 9,420,585 bi-allelic SNPs available in at least two cohorts with different populations. We found that five loci reached *P_int_* < 5e-8, including one locus at 16q23.1 with statistical significance in EUR analysis. It should be noted that three of these five loci are driven by low frequency SNPs. (Table 3; Supplemental Figure 8 for Manhattan plot; Supplemental Figure 9 for regional plots). The lead SNPs in the five loci are intronic variants to *THSD7B*, *SPATA5*, *UBE3D*, *GATA4*, and *WWOX*, respectively. None of the SNPs with *P_int_* < 0.05 at 1 mb region surrounding the five lead SNPs overlapped with SNPs associated with systolic BP (*P* < 1e-5) in the GWAS catalog [43].

**Table 3.**
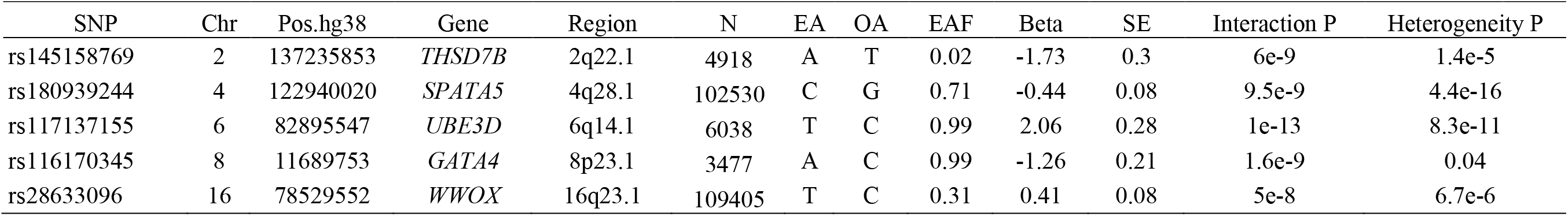
Statistically significant SNPs in *cross-population* meta-analysis of gene-quantitative DASH diet score interaction.

| SNP | Chr | Pos.hg38 | Gene | Region | N | EA | OA | EAF | Beta | SE | Interaction P | Heterogeneity P |
| --- | --- | --- | --- | --- | --- | --- | --- | --- | --- | --- | --- | --- |
| rs145158769 | 2 | 137235853 | *THSD7B* | 2q22.1 | 4918 | A | T | 0.02 | -1.73 | 0.3 | 6e-9 | 1.4e-5 |
| rs180939244 | 4 | 122940020 | *SPATA5* | 4q28.1 | 102530 | C | G | 0.71 | -0.44 | 0.08 | 9.5e-9 | 4.4e-16 |
| rs117137155 | 6 | 82895547 | *UBE3D* | 6q14.1 | 6038 | T | C | 0.99 | 2.06 | 0.28 | 1e-13 | 8.3e-11 |
| rs116170345 | 8 | 11689753 | *GATA4* | 8p23.1 | 3477 | A | C | 0.99 | -1.26 | 0.21 | 1.6e-9 | 0.04 |
| rs28633096 | 16 | 78529552 | *WWOX* | 16q23.1 | 109405 | T | C | 0.31 | 0.41 | 0.08 | 5e-8 | 6.7e-6 |

## Discussion

In this genome-wide interaction analysis in the CHARGE cohorts and the UK biobank, we showed that the association of DASH diet score with systolic BP was modified by multiple SNPs: at three loci in EUR analyses and an additional four loci in cross-population analysis. Our interaction SNP hits are independent of known BP loci from BP-GWAS. Furthermore, at the *MTHFS* locus (15q25.1), we demonstrated that the SNP-DASH diet score interaction may affect systolic BP through regulating levels of DNA methylation at this locus. While limitations exist in this study, our findings provide novel insights into gene-diet interactions in BP with respect to a better understanding of potential mechanisms of BP regulation and more personalized dietary advice.

Hypertension is the leading risk factor for cardiovascular disease [44]. Both environmental and genetic risk factors can lead to hypertension. A recent review summarized gene-lifestyle interactions in relation to hypertension and highlighted several genes that may interact with lifestyle factors to modify the risk of hypertension [45]. For example, several studies investigated interactions between genes and dietary components such as salt, alcohol, and fat intake [19, 46-48]. In a study of 4,414 individuals who participated in the Korean Genome and Epidemiology Study, an interaction was observed between rs3784789 (intronic to *CSK* and upstream to *MIR4513*) and estimated 24-hour urinary sodium to potassium ratio in relation to the risk of hypotension [46]. This SNP was nominally significant in the present interaction analysis, both in the EUR and cross-population analyses (*P* = 0.009 and 0.005, respectively). As also indicated in this previous review study [44], most studies had modest sample sizes, and the observed interactions were often different across studies. In the present study, we also observed that most of the loci with significant interactions had high heterogeneity, specifically the poor replication between observations in the CHARGE cohorts and the UK Biobank (Supplemental Figure 10). This high heterogeneity may be partly due to the diverse food habits that limit the ability of the DASH diet score to consistently reflect overall diet quality across cohorts or different dietary tools used in our participating cohorts. The present study adds novel evidence to the literature regarding interaction between genetic variants and overall diet quality, nonetheless, future analyses with larger sample sizes and better harmonized dietary information are needed to validate our findings.

At 15q25.1, the lead SNP, rs117878928 resides ∼2 kb downstream of long noncoding RNA (*LOC124903536*) and ∼2 kb upstream of a protein coding gene (*MTHFS*). *MTHFS* encodes methenyltetrahydrofolate synthase that catalyzes the conversion of 5-formyltetrahydrofolate to 5,10-methenyltetrahydrofolate. Activation of *MTHFS* may accelerate folate catabolism by modifying folate one-carbon forms, leading to impaired methylation reactions such as DNA methylation [49]. Folate metabolism has been implicated in the risk of hypertension [50, 51], although its role is not fully established. In an earlier study conducted in EUR participants from two participating cohorts of the present study, the Atherosclerosis Risk Communities study (ARIC) and the FHS, an intronic variant (rs6495446) of *MTHFS* has been linked to chronic kidney disease [52], a disease tightly associated with elevated BP. However, rs6495446 is not in LD with rs117878928 (R^2^ = 0.006).

Our observation regarding the enrichment of *cis*-mQTLs in this locus appears to be consistent with the functionality of *MTHFS*. In FHS, SNP rs117878928 is a *cis*-mQTL variant for cg21315874 (*h^2^* = 0.015, *P* = 4.4e-15) [40], which is residing at 5’ untranslated region (UTR) of *ST20*. In the Genetics of DNA Methylation Consortium (GoDMC) [53], rs117878928 is also a *cis*-mQTL variant for other CpGs (e.g., cg22389121, *P*=2.3e-42) at 5’UTR or close the transcription start site of *ST20*. *ST20* is adjacent to *MTHFS*, and it is a tumor suppressing gene involved in several processes such as apoptotic signaling pathway, cellular response to ultraviolet C, and negative regulation of cell growth [54]. *ST20-MTHS* readthrough transcript can be formed through splicing to produce a fusion protein that shares sequence identity from both genes, which are highly expressed in the liver and kidney (https://www.proteinatlas.org) [55]. Overall, our observations indicate that a DNA methylation related mechanism may be relevant to the gene-DASH diet score interaction observed in this region.

The joint analysis of SNP main effects and interaction effects has been shown to be more powerful than the analysis of SNP main effects or interaction effects alone, when the genetic effects are relatively weak and the interaction effects are moderate [36, 56]. Thus, the joint analysis is a promising approach to identify additional loci relevant to traits of interest. In the present study, we compared the joint analysis results with main effect statistics obtained from our study sample and a larger GWAS for systolic BP [15], and we identified several candidate loci for future validation (Supplemental Table 8). For example, the significant 2df test for an intronic SNP (rs140635454) of *NUP93* was driven by its interaction with the DASH diet score. A recent study reported a significant 2df test for rs76976871 at this gene in a sleep-by-gene interaction analysis for high density lipoprotein cholesterol [57]. Although the interaction term was modest in the sleep study [57], our results and theirs combined highlight the importance of conducting a comprehensive joint analysis to facilitate identification of loci that are potentially modified by diet and other lifestyle factors.

There are several limitations in this study that should be discussed. First, the cross-sectional nature of the study does not allow us to account for reverse causation, which might have occurred if individuals with hypertension were advised to change their dietary patterns to help control their blood pressure. The DASH diet score was calculated using different versions of FFQs in the CHARGE cohorts and multiple 24-hour recalls in the UK Biobank. All these dietary assessments tools are based on self-reported dietary intake, which is subjective to both random and systematic bias [58]. In addition, different versions of FFQs have different food lists and different levels of detail, e.g., a 126-item FFQ was used in FHS, while the ARIC Study used a 66-item FFQ; therefore, some DASH diet score components may include a different numbers and types of food items. These combined may partly explain the high heterogeneity regarding gene-DASH diet score interaction across cohorts. The majority of our study participants (91%) were EUR; our cross-population analysis was therefore mainly driven by the EUR participants. Furthermore, the numerous associations detected in the lower quartile dichotomized DASH score are probably due to misclassification, therefore larger sample size to replicate the threshold analysis is necessary.

In conclusion, we demonstrated gene-DASH diet score interaction in several loci, particularly at the *MTHFS* locus (15q25.1). In addition, we showed that DNA methylation may be a relevant mechanism linking gene-diet interaction for systolic BP. Compared to large GWAS, the sample size of the present study is modest; therefore, studies with larger sample size and more diverse populations are needed to validate our findings and to facilitate the measure of inter-individual differences in dietary response, allowing for identification of high risk subgroups who would benefit the most from dietary modifications.

## Data Availability

CHARGE data available through dbGap or by request to specific cohorts. The UK Biobank data is available through UK Biobank data repository. The researchers need a proposal to request access to the data.

## Acknowledgements

**ARIC**

The Atherosclerosis Risk in Communities study (ARIC) has been funded in whole or in part with Federal funds from the National Heart, Lung, and Blood Institute, National Institutes of Health, Department of Health and Human Services (contract numbers HHSN268201700001I, HHSN268201700002I, HHSN268201700003I, HHSN268201700004I and HHSN268201700005I), R01HL087641, R01HL059367 and R01HL086694; National Human Genome Research Institute contract U01HG004402; and National Institutes of Health contract HHSN268200625226C. The authors thank the staff and participants of the ARIC study for their important contributions. Infrastructure was partly supported by Grant Number UL1RR025005, a component of the National Institutes of Health and NIH Roadmap for Medical Research.

**CHS**

This Cardiovascular Health Study (CHS) research was supported by NHLBI contracts HHSN268201200036C, HHSN268200800007C, HHSN268201800001C, N01HC55222, N01HC85079, N01HC85080, N01HC85081, N01HC85082, N01HC85083, N01HC85086, 75N92021D00006; and NHLBI grants U01HL080295, R01HL087652, R01HL105756, R01HL103612, R01HL120393, and U01HL130114 with additional contribution from the National Institute of Neurological Disorders and Stroke (NINDS). Additional support was provided through R01AG023629 from the National Institute on Aging (NIA). A full list of principal CHS investigators and institutions can be found at CHS-NHLBI.org. The provision of genotyping data was supported in part by the National Center for Advancing Translational Sciences, CTSI grant UL1TR001881, and the National Institute of Diabetes and Digestive and Kidney Disease Diabetes Research Center (DRC) grant DK063491 to the Southern California Diabetes Endocrinology Research Center. The content is solely the responsibility of the authors and does not necessarily represent the official views of the National Institutes of Health.

**FHS**

The authors thank the staff and participants of the Framingham Heart Study (FHS) study for their important contributions. The Framingham Heart Study was supported by NIH contracts N01-HC-25195, HHSN268201500001I, and 75N92019D00031.

**NEO**

The authors of the NEO study thank all individuals who participated in the Netherlands Epidemiology in Obesity study, all participating general practitioners for inviting eligible participants and all research nurses for collection of the data. We thank the NEO study group, Petra Noordijk, Pat van Beelen and Ingeborg de Jonge for the coordination, lab and data management of the NEO study. The genotyping in the NEO study was supported by the Centre National de Génotypage (Paris, France), headed by Jean-Francois Deleuze. The NEO study is supported by the participating Departments, the Division and the Board of Directors of the Leiden University Medical Center, and by the Leiden University, Research Profile Area Vascular and Regenerative Medicine. Raymond Noordam was supported by a grant from NHLBI (grant number R01 HL156991).

**Raine Study**

The Raine Study was supported by the National Health and Medical Research Council of Australia [Grant Numbers 572613, 403981, 1059711, 1027449, 1044840, 1021858], the Canadian Institutes of Health Research [Grant Number MOP-82893], and WA Health, Government of Western Australia (WADOH) [Future Health WA G06302]. Funding was also generously provided by Safe Work Australia. The authors are grateful to the Raine Study participants and their families, and to the Raine Study team for cohort coordination and data collection. The authors gratefully acknowledge the NHMRC for their long-term funding to the study over the last 30 years and also the following institutes for providing funding for Core Management of the Raine Study: The University of Western Australia (UWA), Curtin University, Women and Infants Research Foundation, Telethon Kids Institute, Edith Cowan University, Murdoch University, The University of Notre Dame Australia and The Raine Medical Research Foundation. This work was supported by resources provided by the Pawsey Supercomputing Centre with funding from the Australian Government and the Government of Western Australia.

**SP & LVB**

The Multi-Ethnic Cohort and Singapore Population Health Study 2012 studies (from which the Singapore Prospective Study Program [SP] and Living Biobank [LVB] are drawn from are supported by individual research and clinical scientist award schemes from the National Medical Research Council (NMRC) and the Biomedical Research Council (BMRC) of Singapore, the Singapore Ministry of Health, National University of Singapore and National University Health System, Singapore.

**WHI**

Women’s Health Initiative (WHI) funded by the National Heart, Lung, and Blood Institute, National Institutes of Health, U.S. Department of Health and Human Services through contracts HHSN268201600018C, HHSN268201600001C, HHSN268201600002C, HHSN268201600003C, and HHSN268201600004C. The authors thank the WHI investigators and staff for their dedication, and the study participants for making the program possible. A full listing of WHI investigators can be found at: https://www-whi-org.s3.us-west-2.amazonaws.com/wp-content/uploads/WHI-Investigator-Long-List.pdf. Additional support to NF was provided by NIH DK117445 and MD012765.

**UK Biobank**

UKB (<u>United Kingdom Biobank</u>): The authors thank the staff and participants of the UK Biobank study for their important contributions. This research has been conducted using the UK Biobank Resource under the UK Biobank project number 56735 and application title “Interaction of Diet Quality Scores and Genetic Variants in Relation to Cardiometabolic Traits”.

## Disclosed Funding and sources of support

JM: Supported by K22-HL-135075; DCR: R01 HL118305 and R01 HL156991; PBM and HRW acknowledge the support of the NIHR Biomedical Research Centre, Queen Mary University of London; NR funding: R01-MD012765, R01-DK117445, R01-HL163972; This project was partly supported by the Intramural Research Program of the National Human Genome Research Institute of the National Institutes of Health through the Center for Research on Genomics and Global Health (CRGGH).

---

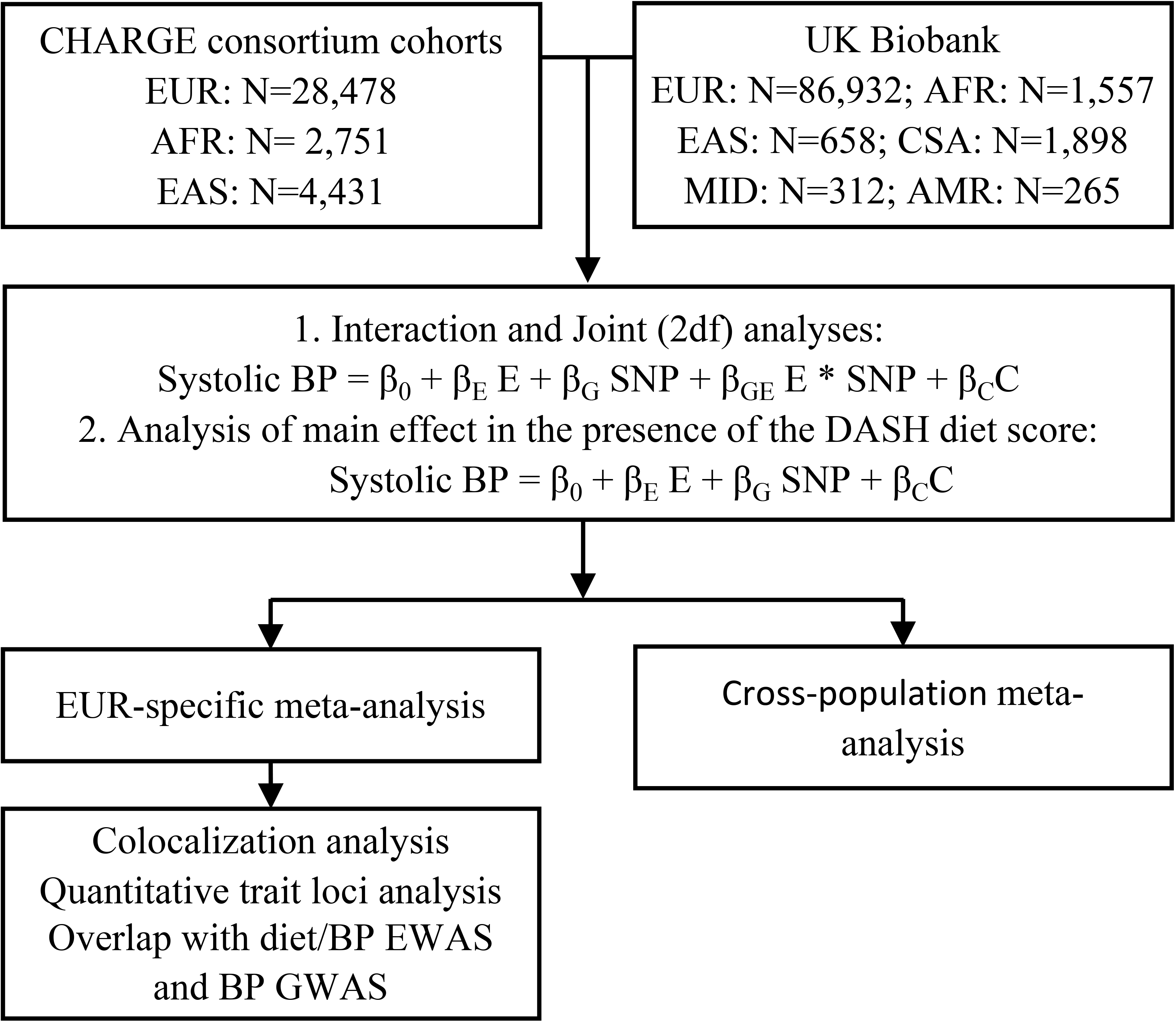

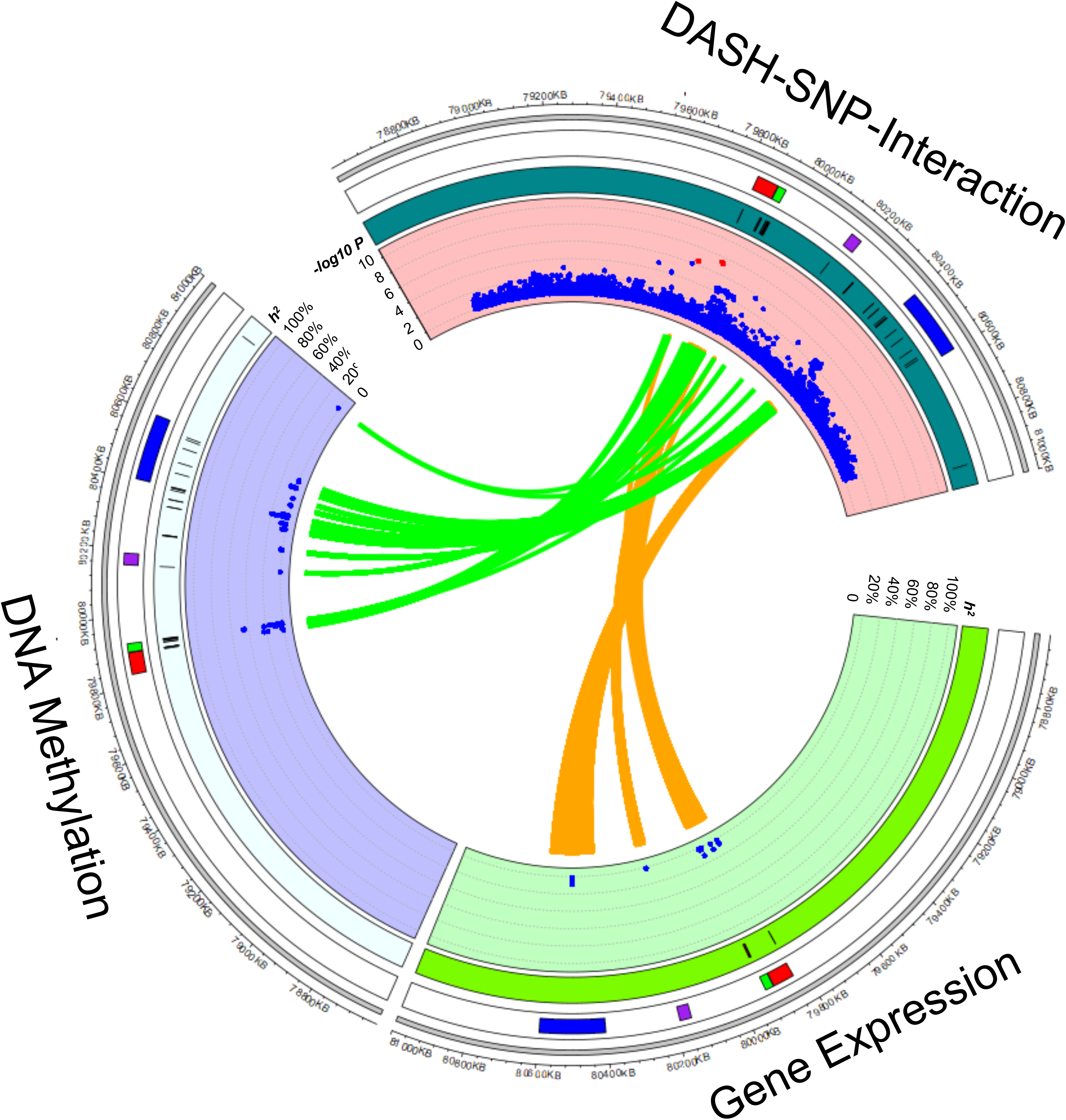

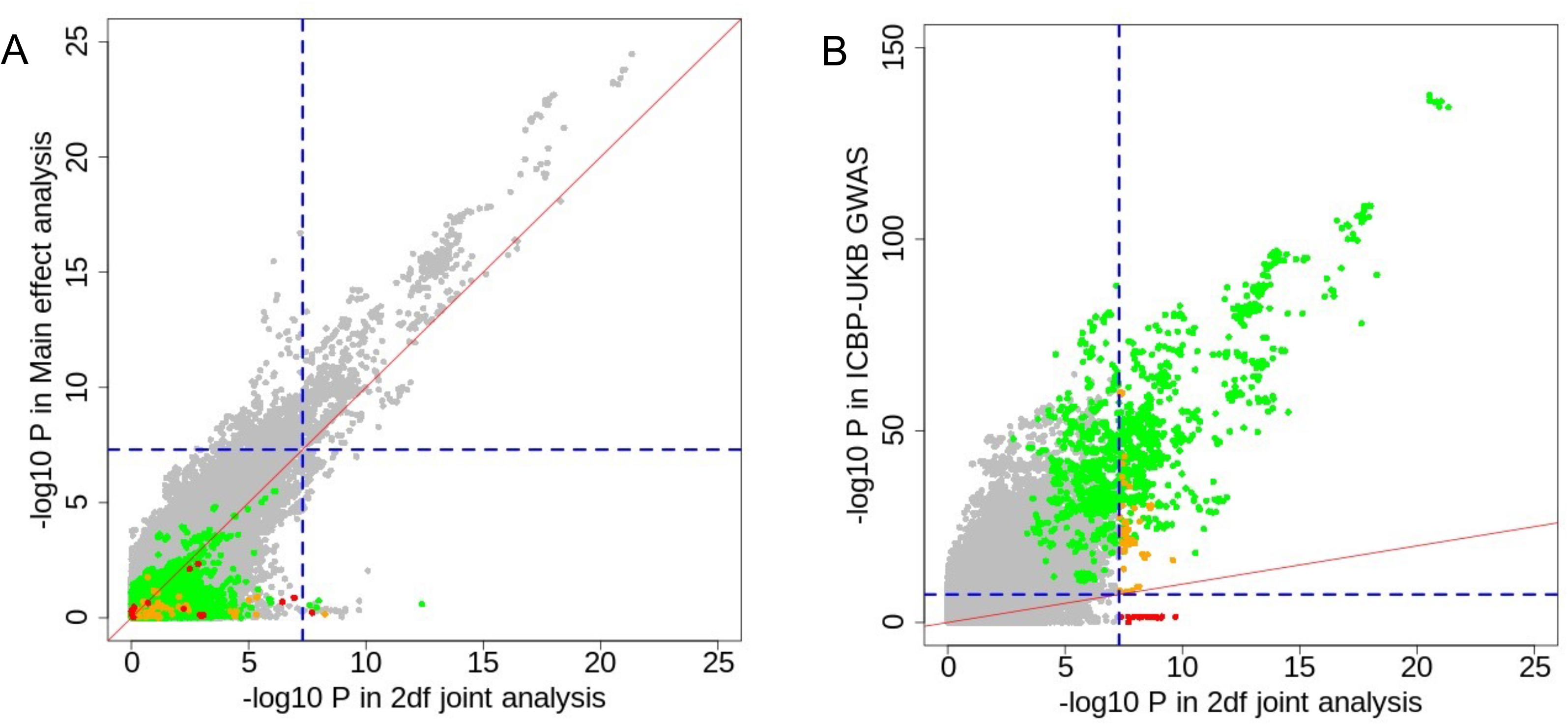

## Notes

### Competing Interest Statement

The authors have declared no competing interest.

### Author Declarations

The present study protocol was approved by the Tufts University Institutional Review Board.

